# Outpatient Portal Use and Blood Pressure Management during Pregnancy

**DOI:** 10.1101/2024.10.21.24315766

**Authors:** Athena Stamos, Naleef Fareed

**Author notes:** Naleef Fareed, PhD, MBA, 460 Medical Center Drive, Columbus, OH 43210, United States. **Declarations**. **Competing Interest Statement:** The authors have declared no competing interest. **Ethics approval and consent to participate:** Retrospective data were obtained for patient encounters from the Academic Medical Center Information Warehouse in accordance with the Honest Broker protocol. This study is recognized by The Ohio State University Institutional Review Board to use de-identified data, and formal Institutional Review Board approval was waived. **Funding:** Time for the work of NF was partially funded by HS028822 from the Agency for Healthcare Research and Quality. The content is solely the responsibility of the authors and does not necessarily represent the official views of the National Institutes of Health or the Agency for Healthcare Research and Quality. **Contributors:** NF provided oversight for the study design and analysis. AS and NF conducted the analysis. AS and NF wrote the manuscript. All authors revised the manuscript for relevant scientific content and approved the final version of the manuscript.

## Abstract

We investigated the association between systole and diastole, and outpatient portal use during pregnancy. We used electronic and administrative data from our academic medical center. We categorized patients into two groups: (<140 mm Hg; <90 mm Hg), and out-of-range (≥140 mm Hg, ≥ 90 mm Hg). Random effects linear regression models examined the association between mean trimester blood pressure (BP) levels and portal use, adjusting for covariates. As portal use increased, both systole and diastole levels decreased for the out-of-range group. These differences were statistically significant for patients who were initially out-of-range. For the in-range group, systole and diastole levels were stable as portal use increased. Results provide evidence to support a relationship between outpatient portal use and BP outcomes during pregnancy. More research is needed to expand on our findings, especially those focused on the implementation and design of outpatient portals for pregnancy.

## Introduction

Outpatient portals can improve health outcomes by facilitating communication between patients and medical providers, encouraging patient engagement with their own care, and promoting patient self-management through digital tools [1-4]. In particular, outpatient portal use in certain clinical contexts has been linked to better results for patients with chronic conditions, including diabetes, hypertension, and depression [5-7]. Individuals with chronic conditions can easily manage their visits, access lab results, and communicate with their health providers using outpatient portals.

The evidence base, however, on the relationship and the mechanisms between patient portal use and better health outcomes is limited and, at times, ambiguous. The few studies on patient portal use have primarily examined user characteristics and user satisfaction [8]. Outpatient portals allow patients with type 2 diabetes to self-manage their blood pressure (BP) [9] which can lower BP levels [10]. High BP levels can have a negative impact on pregnancy outcomes [11-13]. A leading cause of maternal morbidity and mortality is hypertensive disorders of pregnancy, which include chronic hypertension, gestational hypertension, and preeclampsia/eclampsia [14,15]. Hypertensive disorders of pregnancy can have long-term effects on the mother such as heart failure or chronic kidney disease [14].

Prior research have investigated outpatient portals among non-pregnant adults, with limited information on pregnant individuals [16-18]. There is some evidence to suggest that outpatient portal use during pregnancy can improve health outcomes [19-21]. Ukoha and colleagues confirmed that the current literature lacks examination of the effects of outpatient portal use with pregnancy outcomes. Moreover, studies that explore outpatient portal use on clinical outcomes require a substantial sample size to provide appropriate statistical power.

Outpatient portals, like various other digital health tools, can theoretically provide pregnant individuals with information and resources about their health between and during clinical encounters to support beneficial health behaviors, which can impact maternal and infant health outcomes [18,19]. There is research to indicate the presence of differences in outpatient portal use among pregnant individuals [22].

Managing systole and diastole with the help of outpatient portal technology could help lower the risk of pregnant individuals of potential complications with hypertensive disorders of pregnancy. Very few studies have examined the effects of outpatient portal use on BP management during pregnancy. We aimed to bridge this gap in the current literature. The objective of our study was to examine the association between systole and diastole, and outpatient portal use among pregnant individuals.

## Methods

### Data collection

We conducted a retrospective study of pregnant individuals seen at our Academic Medical Center (AMC). Individuals were included if they were pregnant, at least 18 years old or older, received prenatal care at our AMC, and had a registered portal account between 1/1/2016 and 8/1/2020. This period reflects a stable time period of outpatient portal system use at our AMC. We restrict our primary analytical sample to the first pregnancy episode at the AMC to ensure a comparable sample that is not differentially influenced by experience gained in the use of the portal or changes in BP measurements due to multiple pregnancies. The dataset was structured as unbalanced panels for our primary regression analyses, meaning patients may not have a BP outcome recorded every trimester. Before applying our exclusion criteria, we had 17,411 patients. The majority of patients that were excluded were due to missing blood pressure readings and due to no portal use. Our unbalanced panel had 8,870 patients and 23,159 observations.

### Primary outcome variables

The primary outcomes of interest were systole and diastole measurements collected between the time of first encounter during the pregnancy until delivery. BP outcome measures were from in person clinical encounters. Our analytical dataset included individuals who had at least one BP measurement in the first or second trimester and one in the third trimester. The outcome measures were treated as continuous variables, and we computed geometric means within trimesters if multiple readings were collected.

### Patient portal

Our AMC’s MyChart [23] portal is a secure electronic system that is linked to a patient’s health record. This free and personalized portal facilitates patient provider communication through features such as secure electronic messaging, appointment scheduling, availability of lab results, and billing. We defined overall outpatient portal use as the number of times (i.e., sessions) the portal was used by a patient with a registered account in a trimester. We calculated the frequency of use of each of nine portal features (messaging, visits, my record, medical tools, billing, resources, proxy, preferences, and custom, which includes use of miscellaneous features) as well as the overall frequency of use of all nine features within a trimester. For our primary analysis, we excluded patients who used the portal less than three times in a trimester, which represented the bottom 25^th^ percentile of portal sessions in our data. Details on the data processing can be found elsewhere [22].

### Primary exposure variable

The primary exposure of interest was the interaction between outpatient portal use and an out-of-range flag for BP measures. We grouped individuals into similar groups: in-range (<140 mm Hg; <90 mm Hg) and out-of-range (≥140 mm Hg, ≥ 90 mm Hg). These cut offs were based on guidelines from the American College of Obstetrics and Gynecology [24].

### Covariates

Patient level covariates included demographic factors such as racioethnicity (“non-Hispanic white”, “non-Hispanic Black”, “Hispanic” and “Other racioethnicity groups”), age at first prenatal visit, insurance status (“Commercial,” “Medicaid,” and “Other”), and pregnancy associated clinical risk (“Normal” vs “High-Risk”). Individuals were classified as having high pregnancy related risk if they had diabetes, high blood pressure, preeclampsia, advanced maternal age, multiple pregnancies, history of premature birth, genetic conditions, or any condition requiring high-risk care or fetal treatment. We included a community-based composite measure of opportunity called the Ohio Opportunity Index (OOI) to account for any associated socio-demographic and neighborhood differences. The community social determinants of health (cSDoH) score, as measured by the OOI, describes the relative deprivation/opportunity for health and well-being of adults based on their ZIP Codes across the state of Ohio using national and state-specific measures. Higher scores indicate relatively greater opportunity or lower deprivation. For further details about this measure refer to Fareed and colleagues [25]. We also included an area-level measure for neighborhood household internet connectivity using the National Neighborhood Data Archive: Internet Access by Census Tract and ZIP Code Tabulation Area, United States, 2015-2019 [26]. This operationalized as a binary variable with 1 indicating neighborhood access to DSL, cable, or other highspeed internet, and 0 indicating a lack of access. These covariates were selected because these characteristics may confound the relationship between portal use and health outcomes during pregnancy, supported also by previous literature that examined associations between portal use and BP outcomes [8,27].

### Ethics

Retrospective data were obtained for patient encounters from AMC’s Information Warehouse (IW) in accordance with the Honest Broker protocol; this protocol is recognized by the Institutional Review Board (IRB) to use de-identified data without requiring formal IRB approval.

### Statistical Analysis

Patient data were collected at the trimester level. Our preliminary inspection suggested skewness in portal use data, thus we transformed them logarithmically for further analyses. We report descriptive statistics for our analytical dataset. Our main analysis estimates the association of the logged sum of overall portal use (or logged sum of each portal function used) with each of the BP outcomes using multivariable random-effects linear regression models. We created an interaction of the primary predictor with the out-of-range variable for a BP outcome (1= out-of-range, 0=in-range). We provide a general specification of our empirical model below:

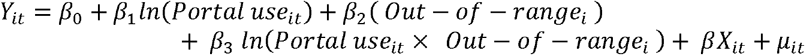

For this model, *i* indexes the patient and *t* indexes the pregnancy trimester (t =1, 2, 3). *Y*_*it*_ is the BP measure of interest. *Portal use*_*it*_ refers to the log of the total number of times the portal feature(s) were used during a trimester. *Out* – *of* – *range*_*i*_ is a binary variable for a patient who is out of range in the first or second trimester (1= out-of-range, 0 otherwise). The interaction term *Portal use*_*it*_ × *Out* – *of* – *range*_*i*_ is set equal to the log of the total number of times the portal features(s) were used for individuals that were out-of-range or zero otherwise. *β X*_*it*_ is a vector of covariates (racioethnicity, age, insurance status, clinical risk, households with internet availability, and cSDoH score). The term *μ*_*it*_ represents random intercepts. We computed cluster-robust standard errors by patient to make standard errors robust. We estimate the adjusted predicted BP outcomes by portal use for individuals out-of-range and in-range to explore within and between changes in the outcomes. We excluded any cases with missing information for our regression model (complete case analysis). All analyses were conducted using Stata MP 18.0. We report our study findings following STROBE guidelines [28]. Our sample size is adequately powered to detect a small effect size, with moderate interclass correlation, α of 0.05, and power of 0.80.

### Sensitivity Analysis

For our sensitivity analysis, we included multiple pregnancies and ran a similar model specification as our primary analysis to explore robustness of our estimates. We also ran the random effects model by trimesters and racioethnicity to examine possible differences in our results. Lastly, we performed our main analysis with a balanced panel (i.e. patients who had BP measurements in all three trimesters of their first pregnancy).

## Results

### Patient Characteristics

Table 1 presents characteristics of patients in the unbalanced panels. There were 8,870 patients with a total of 23,159 observations, as each patient had BP data from at least two trimesters. The average systole and diastole values were 116 mm Hg and 69 mm Hg respectively. Approximately 2% and 1% of our observations had out-of-range values for systole and diastole, respectively. The average age of individuals in this sample was 30 years old, 69% identified with non-Hispanic white racioethnicity, over 70% had commercial insurance, and about 20% had high pregnancy related risks. The average cSDoH score was 81, which was also lower than the state of Ohio’s mean.

**Table 1.** Characteristics of the analytical sample consisting only of first pregnancy episodes.

| Variable | BP<br>N=23,159 |
| --- | --- |
| BP outcome (mean, SD) |  |
| Systole: Overall | 115.80 ($\pm 9.63$) |
| Systole: In-range | 115.28 ($\pm 8.88$) |
| Systole: Out-of-range | 139.02 ($\pm 12.69$) |
| Diastole: Overall | 69.47 ($\pm 7.14$) |
| Diastole: In-range | 69.24 ($\pm 6.81$) |
| Diastole: Out-of-range | 87.54 ($\pm 9.20$) |
| Age in years at initial encounter (mean, SD) | 30 ($\pm 5$) |
| Racioethnicity (n, %) |  |
| Non-Hispanic white | 16,013 (69.14%) |
| Non-Hispanic Black | 3,714 (16.04%) |
| Hispanic | 735 (3.17%) |
| Other Racioethnicity | 2,697 (11.65%) |
| Percentage of households with Internet availability (mean, SD) | 87.45% ($\pm 8.47\%$) |
| Insurance status (n, %) |  |
| Commercial | 16,406 (70.84%) |
| Medicaid | 4,443 (19.18%) |
| Other/Self-Pay | 2,310 (9.97%) |
| Clinical Risk (n, %) |  |
| Normal | 18,548 (80.09%) |
| High | 4,611 (19.91%) |
| Community SDoH (mean, SD) | 81.44 ($\pm 11.55$) |
| Individuals out-of-range (n, %) |  |
| Systole | 507 (2.19%) |
| Diastole | 288 (1.24%) |
| Portal use by trimester (mean, SD) |  |
| 1<sup>st</sup> Trimester |  |
| Sum of portal use | 14.86 ($\pm 11.65$) |
| Logged sum of portal use | 2.45 ($\pm 0.71$) |
| 2<sup>nd</sup> Trimester |  |
| Sum of portal use | 30.60 ($\pm 23.46$) |
| Logged sum of portal use | 3.13 ($\pm 0.80$) |
| 3<sup>rd</sup> Trimester |  |
| Sum of portal use | 34.42 ($\pm 27.18$) |
| Logged sum of portal use | 3.23 ($\pm 0.82$) |

Note: Systole and Diastole levels measured in mm Hg; Age at initial encounter measured in years; BP=Blood pressure; SDoH=Social Determinants of Health.

### General effects between BP outcomes and portal use

The estimates indicated significant differences in mean trimester systole and diastole levels among out-of-range and in-range individuals based on their mean trimester portal use. Both systole and diastole levels were lower in individuals of non-white racioethnicity, and higher in those living in areas with lower cSDoH scores. Individuals with high pregnancy associated risks also had higher systole and diastole levels.

We examined the association between mean trimester systole and diastole levels and the mean trimester use of each portal feature. The results suggested that the use of three popular features: visits, myRecords, and messaging contributed the most to the overall reduction in BP (results available upon request).

### Within group comparison

Figure 1 illustrates the mean trimester predicted values for the in-range and out-of-range groups by each BP outcome. We found greater reductions associated with mean trimester portal use in the out-of-range group for both BP outcomes. For the out-of-range group, the differences in mean trimester predicted systole and diastole between the lowest and highest mean trimester overall portal use levels were 31.54 mm Hg (95% CI: 30.15, 32.93) and 18.23 mm Hg (95% CI: 16.68, 19.78), respectively. Systole and diastole levels increased slightly for the in-range group as overall portal use increased.

**Figure 1.**
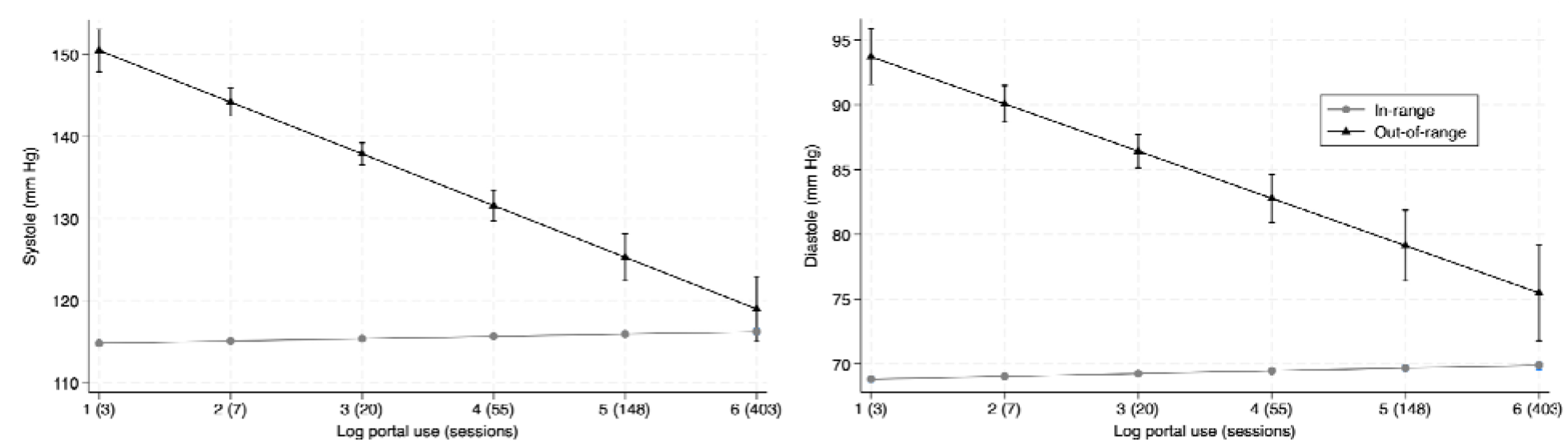
Mean trimester predicted systole and diastole based on log sum portal in the unbalanced panels (n=8,87 patients N= 23,159 observations). The average trimester portal use in our dataset ranged from 0 and 14 sessions per week over a trimester.

### Between group comparison

Figure 1 illustrates the differences in the changes between the in-range and out-of-range groups for the BP outcomes across different levels of mean portal use per trimester. The lowest portal users had mean trimester differences of 35.73 mm Hg (95% CI: 33.46, 38.01) and 24.89 mm Hg (95% CI: 22.99, 26.78) for systole and diastole, respectively. These differences decreased as mean trimester overall portal use increased. However, systole was not statistically significant between the in-range and out-of-range groups at the highest portal use level.

### Sensitivity analysis

We conducted a sensitivity analysis keeping multiple pregnancies among individuals over time in the model. There was a reduction of 1.17 mm Hg in diastole for individuals in the out-of-range group at the highest level of portal use compared to our primary analysis. We also examined the differences between the in-range and out-of-range groups by trimester and across Non-Hispanic (NH) White and NH Black racioethnic populations (results not shown). We found similar reductions in BP based on higher outpatient portal use as the reported predicted means from our primary analysis. Additionally, we replicated our primary analysis using the balanced panel dataset. The out-of-range group had a lower mean trimester predicted systole compared to the in-range group for the highest portal use. There was a difference of 2.90 mm Hg at the highest portal use between the in-range and out-of-range groups for systole.

## Discussion

Our primary finding suggests that outpatient portal use can affect two critical BP outcomes (systole and diastole) during pregnancy. These effects were especially significant for those whose initial measurements were out-of-range. The American Heart Association defines Stage 2 hypertension as systole greater than 140 mm Hg or diastole at least 90 mm Hg [29]. Systole less than 120 mm Hg and diastole less than 80 mm Hg are considered normal [29]. As overall portal use increased, the out-of-range group improved from Stage 2 hypertension to normal ranges for both systole and diastole. Based on our results, after approximately 10-20 sessions on the outpatient portal per trimester, pregnant individuals achieve a BP range considered safe according to the American College of Obstetrics and Gynecology [24]. Given the association between these BP measures and maternal morbidity as well as other pregnancy complications, these significant changes found in our analyses may be associated with improved maternal and infant outcomes. Previous studies have found hypertensive disorder-related Severe Maternal Morbidity (SMM) greatly linked to bad health outcomes [30,31]. Hypertensive disorders can affect placental function and fetal growth, increasing the risk of SMM [31]. Additionally, a reduction of 10 mm Hg in systole is associated with a 41% decrease in stroke risk and a 22% decrease in ischemic heart disease [32].

Our findings indicated that the three portal functions: myRecords, visits, and messaging contributed the most to the overall reduction in mean trimester BP outcomes. MyRecords allows patients to access their medical data such as lab results and health conditions [22]. The visits feature helps patients manage their appointments and the messaging function allows communication between patients and providers [22]. The use of these three functions may be explained by the Theory of Planned Behavior (TPB; [33]). These functions may support clinically meaningful activities during pregnancy that address TPB elements (i.e., attitudes, subjective norms, and perceived control for engaging in a health behavior) that may be subsequently linked to improved health outcomes, such as BP levels. For example, having summaries of one’s medications, test results, and plan of care may provide more information that empowers pregnant individuals to have conversations with their care team about factors that can influence their BP and other health outcomes, which may concomitantly transform patient attitudes, normative beliefs, and perceived control about their health behavior.

A prior study found that pregnant individuals mainly used the myRecords, visits, and messaging functions [22]; however, this study did not explore linkages to pregnancy health outcomes. Another study by Holder and colleagues did not find significant differences in systole levels during pregnancy among outpatient portal users [34]. Based on our results, there is indication that a more significant reduction in BP may be achievable with the use of outpatient portals among individuals with uncontrolled physiological outcomes. For example, we found that there was a significant reduction in out-of-range systole among the out-of-range group who frequently used the outpatient portal, especially the visits and messaging functions. Given the importance of using specific portal functions, it is important to recognize variability in the implementation and user-centered design of outpatient portals, especially across the settings that have studied portal use with health outcomes [2], which may have been previously overlooked.

Additional research is needed to investigate the interplay between socio-demographic characteristics of pregnant individuals and their outpatient portal use with their health outcomes. Future studies can benefit from the examination of other physiological outcomes, such as gestational weight gain in pregnant individuals. This could provide a deeper understanding of how outpatient portals affect maternal and infant health. The utility of incorporating patient generated health data in digital health tools during pregnancy is increasingly being explored as an important path through which tools such as the outpatient portal can improve health behavior and outcomes [35]. However, the power of gaining information such as BP readings via outpatient portals will need further examination to learn about barriers and facilitators related to patient and care team acceptance and use of such information.

## Limitations

There are limitations to our study. Log file data contains noise from system measurement such as incomplete portal use data, inactive patients, and portal use outside of established timeframes, but we have a well-developed approach to processing this data [22,36]. BP were not prospectively collected for this study and measurement of BP may not be reliable due to human or computer error, but we incorporated values using multiple measures and use geometric mean when available to account for possible outliers. Our study is limited to our AMC; however, the MyChart instance at our AMC uses functions that are ubiquitous to other EPIC instances nation-wide and this could make our results generalizable. We were also unable to track behavioral changes or whether a patient took any medications such as treatments for blood pressure during their pregnancy. However, we did control for patients with clinical risk. These limitations are similar to those of many other studies using EHR data.

## Conclusion

Our study is one of the first studies to have examined the effect of outpatient portal use on BP outcomes during pregnancy. Systole and diastole levels were stable as overall portal use increased for the in-range group. For the out-of-range group, both systole and diastole levels decreased as portal use increased. Although the significant effects we found were per trimester over the pregnancy, there may be potential for outpatient portals to improve maternal and infant outcomes in the long-term, including the postpartum period – acknowledged as critical time point for mothers. More research is needed to learn about how outpatient portals can be designed around pregnancy and whether centering design around the pregnancy journey can enhance health behaviors and maternal and infant outcomes.

## Data Availability

The data is specific to the Academic Medical Center and cannot be made public.

